# Associations between Monocyte-to-High Density Lipoprotein Ratio and abdominal aortic calcification: Results of a nationwide survey

**DOI:** 10.1101/2024.02.27.24303475

**Authors:** Wei Ran

## Abstract

**Background:** Abdominal aortic calcification (AAC), a critical marker for cardiovascular disease, is strongly correlated with metabolic disorders like diabetes and hypertension. Recent studies have highlighted the Monocyte-to-High Density Lipoprotein Ratio (MHR) as a potential biomarker for assessing the risk of atherosclerosis and cardiovascular diseases.

**Methods and Results:** This cross-sectional study utilized data from the National Health and Nutrition Examination Surveys (NHANES) 2013-2014, focusing on U.S. adults aged 40 years and above. A total of 3017 participants were included, with AAC evaluated using dual-energy X-ray absorptiometry (DXA) scans, and the severity of AAC quantified based on the Kauppila score system. MHR was investigated for its association with AAC severity, employing multiple linear and multivariable logistic regression models to explore the relationship dynamics. After adjusting for potential confounders, including age, sex, race, socioeconomic factors, and other health-related variables, the analysis revealed a significant association between higher MHR levels and increased AAC scores. Participants with elevated MHR exhibited a greater prevalence and severity of AAC.

**Conclusions:** The study demonstrates a significant association between elevated MHR and the prevalence and severity of AAC, suggesting the utility of MHR as a predictive biomarker for cardiovascular risk assessment. These findings advocate for the inclusion of MHR in cardiovascular disease management and risk stratification protocols.

## Introduction

Vascular calcification (VC) is a pathology characterized by ectopic calcification in the vessel walls of muscular or elastic arteries[1], commonly observed in patients with diabetes[2], chronic kidney disease (CKD)[3], hypertension, osteoporosis[4], etc. The abdominal aorta is a frequent site of VC. Abdominal aortic calcification (AAC) has been widely acknowledged as an independent predictor of morbidity and mortality from cardiovascular events and as a reliable marker of both subclinical atherosclerotic disease and arteriosclerosis[5–7]. With increasing AAC severity, the risk of fatal cardiovascular events and mortality significantly escalates[8, 9]. Consequently, AAC has garnered increasing attention in recent years.

Monocyte-to-high density lipoprotein (HDL) ratio (MHR) is a novel composite predictor that reflects the balance between the inflammatory and oxidative stress of monocytes and HDL[10]. MHR has been proposed as an indicator of atherosclerotic severity, assessing the degree of dyslipidemia and inflammation[11]. MHR’s predictive ability for clinical outcomes may surpass that of independent monocyte count and HDL concentration[10]. Previous studies have underscored its value in predicting a variety of cardiovascular diseases[12–15].Related studies have confirmed its pivotal role in predicting stroke, congenital heart defects (CHD), and numerous other atherosclerotic diseases, establishing it as a novel prognostic marker for cardiovascular diseases (CVDs). However, research on MHR and CHD in the general population remains nascent.

Consequently, this research aims to explore the relationship between MHR and AAC in US adults, endeavoring to mitigate the interference of confounding factors, utilizing the National Health and Nutrition Examination Surveys (NHANES) database. It seeks to offer novel approaches and methods for the prevention and treatment of AAC.

## Materials and Methods

### Study population

We obtained data from the NHANES, a cross-sectional study designed to evaluate the nutrition and health status of the non-institutionalized U.S. population on a repeating 2-year cycle, conducted by the National Center for Health Statistics (NCHS)[16]. All NHANES data are publicly available at https://www.cdc.gov/nchs/nhanes/.

Our study utilized data from NHANES 2013 to 2014, as this was the only cycle to include comprehensive data on abdominal artery calcification and dietary inflammatory index. Given that data on AAC scores were unavailable for participants aged under 40 years (due to their exclusion from the DXA scan in the NHANES 2013–2014 cycle), we included only participants aged 40 years and above with complete data on AAC scores and DII in our analysis. Initially, 10,175 individuals were enrolled. After excluding participants lacking AAC data (n = 7035) and those missing HMR data (n = 123), a total of 3017 subjects aged 40 years and above were included in our final analysis (Figure 1).

**Figure 1:**
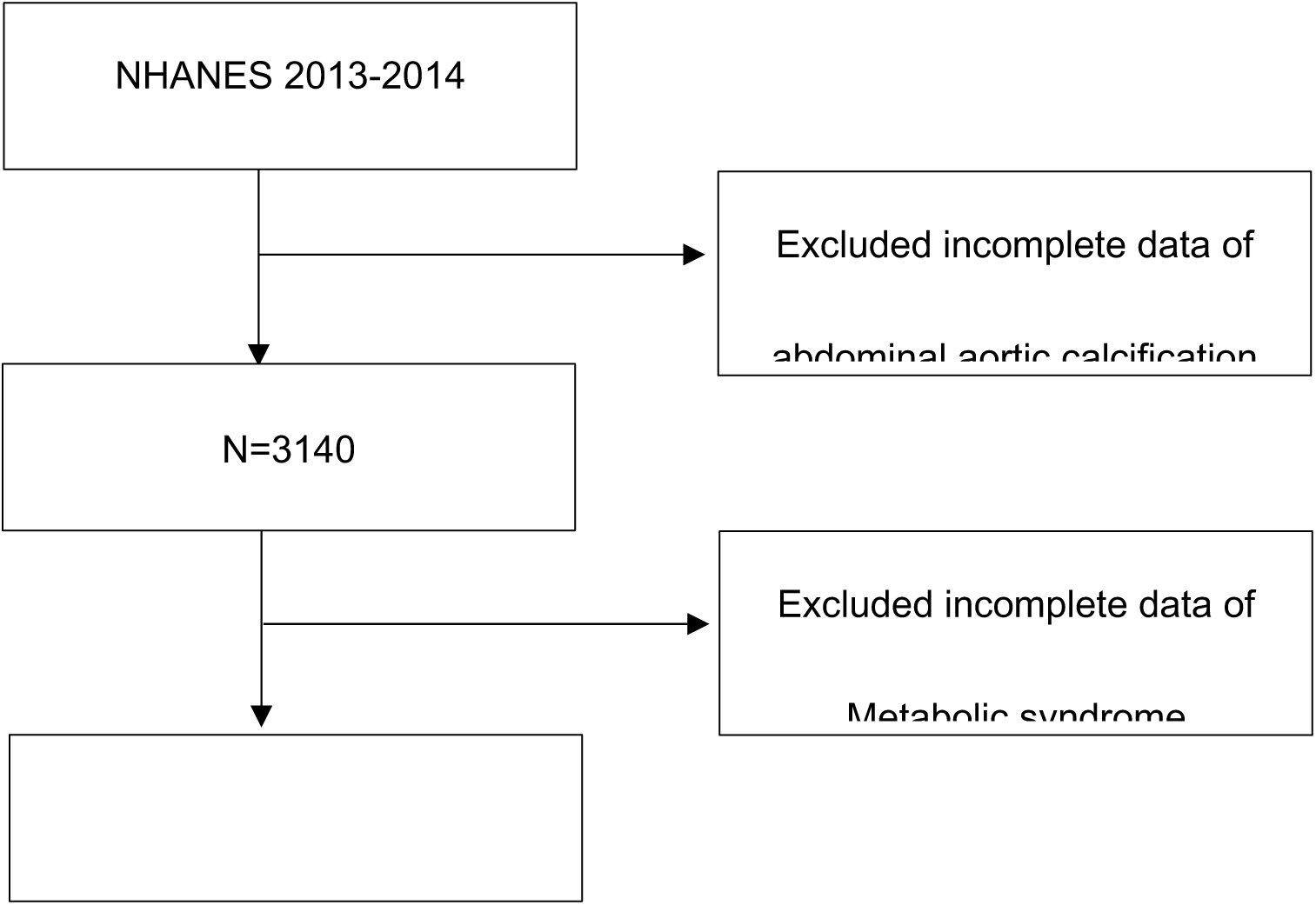
Flow chart of participants selection. NHANES, National Health and Nutrition Examination Survey.

### Exposure and Outcome Definitions

Blood specimens were collected in the morning after a 9-hour fast, following established protocols. The Beckman Coulter MAXM was used to provide complete blood counts for the blood specimens. Monocytes were isolated from white blood cells through three simultaneous measurements: laser light scatter, high-frequency conductivity, and individual cell volume. The concentration of HDL was determined using either direct precipitation or immunoassay[17]. MHR was calculated by dividing the monocyte count (10^^3^ cells/μL) by the HDL concentration (mg/dL).

AAC prevalence, and severe AAC prevalence were identified as outcome variables. The AAC score was quantified through the evaluation of lateral lumbar spine images acquired via DXA (Densitometer Discovery A, Hologic, Marlborough, MA, USA), conducted by trained NHANES-affiliated staff as a single measurement in a mobile examination center (MEC). The total AAC score for each participant was determined strictly according to the Kauppila scoring method, ranging from 0 to 24, by professional technologists. This method has been widely reported and utilized in assessing the severity of calcified vessels[18]. The higher the AAC score, the more severe the calcification condition of the abdominal aorta. Furthermore, AAC is defined as a total AAC score greater than 0, while severe AAC is defined as a total AAC score exceeding 6, consistent with the significance of calcified abdominal aorta as established in previous studies[19, 20].

### Covariates

Continuous variables in our study included age (years), body mass index (BMI, kg/m^2), systolic blood pressure (SBP, mmHg), diastolic blood pressure (DBP, mmHg), and serum creatinine (mg/dL). Categorical variables comprised gender, race, education level, ratio of family income to poverty (RIP), hypertension, diabetes, and smoking status. The detailed measurement processes for these variables are publicly available at www.cdc.gov/nchs/nhanes/.

Statistical analysis was conducted in accordance with CDC guidelines, with appropriate NHANES sampling weights applied to account for the complex multistage cluster survey design utilized in the analysis.

### Statistical Analysis

Continuous variables were reported as mean ± standard error (SE), while categorical variables as percentages. A weighted Student’s t-test and a weighted chi-square test were utilized for continuous and categorical variables, respectively, to assess differences in MHR-tertile groups. MHR, exhibiting a right-skewed distribution, was log-transformed (base 2) for regression analysis. Pearson correlation analysis was conducted to examine the relationship between covariates and MHR. Multivariate logistic regression models were utilized to investigate the independent relationship between MHR and ACC, incorporating AAC score, AAC, and severe AAC, across three distinct models. Adjustments in Model 2 included gender, age, and race, while Model 3 additionally accounted for education level, BMI, SBP, DBP, serum creatinine, ALT, AST, serum calcium, serum phosphorus, total cholesterol, hypertension, and diabetes. Furthermore, for sensitivity analysis, MHR was transformed from a continuous to a categorical variable (tertiles). Stratified multivariate regression analysis was conducted for subgroup analysis, stratified by gender, age, BMI, hypertension, and diabetes. Additionally, an interaction term was introduced to examine heterogeneity in associations among subgroups. A p-value < 0.05 was deemed statistically significant. Analyses were executed using Empower software and R version 4.1.2.

## Result

### Baseline Characteristics of the Enrolled Participants

The basic characteristics of participants, segmented into included and excluded groups, are detailed in Figure 1. Our study incorporated 3,017 middle-aged and elderly adults. Table 1 summarizes the baseline characteristics across MHR-Tertile 1, MHR-Tertile2, and MHR-Tertile3 categories, revealing significant disparities across various demographics and health metrics including sex, race and ethnicity, education levels, Poverty Income Ratio (PIR), estimated Serum Creatinine (Scr), Body Mass Index (BMI), Systolic Blood Pressure (SBP), Hemoglobin A1c (HbA1c), and AAC Scores (all p-values < 0.05). Notably, the high-MHR group was predominantly male, with a higher PIR, and exhibited elevated rates of hypertension and diabetes, increased estimated glomerular filtration rate, and elevated levels of Scr, Total Cholesterol (TC), and SBP, alongside lower phosphorus levels.

**Table 1:** Baseline characteristics of participants according to bone mineral density.

| MHO tertile | Tertile 1 | Tertile 2 | Tertile 3 | P-value* |
| --- | --- | --- | --- | --- |
| Age(years) | 57.48 ± 11.13 | 57.25 ± 11.50 | 57.51 ± 11.91 | 0.8657 |
| Gender (%) |  |  |  | <0.001 |
| Male | 27.56 | 49.75 | 66.73 |  |
| Female | 72.44 | 50.25 | 33.27 |  |
| Race (%) |  |  |  | 0.0054 |
| Mexican American | 6.02 | 8.21 | 6.92 |  |
| Other Hispanic | 4.91 | 4.24 | 4.66 |  |
| Non-Hispanic White | 68.09 | 72.13 | 74.07 |  |
| Non-Hispanic Black | 12.28 | 8.68 | 8.53 |  |
| Other Race | 8.7 | 6.73 | 5.82 |  |
| Education level (%) |  |  |  | <0.0001 |
| Less than high school | 12.75 | 13.11 | 19.39 |  |
| High school or GED | 19.22 | 20.73 | 25.24 |  |
| Above high school | 68.04 | 66.15 | 55.37 |  |
| Marital status (%) | 68.15 | 68.14 | 71.37 | 0.5688 |
| PIR levels | 3.29 ± 1.67 | 3.11 ± 1.68 | 2.92 ± 1.61 |  |
| Smoke |  |  |  | <0.0001 |
| Never | 7.09 | 9.99 | 17.22 | <0.0001 |
| Former | 5.44 | 4.81 | 6.67 |  |
| Current | 87.46 | 85.2 | 76.12 |  |
| Alcohol |  |  |  |  |
| Never | 61.93 | 56.31 | 44.99 | <0.0001 |
| Former | 25.59 | 28.52 | 30.06 |  |
| Current | 12.48 | 15.16 | 24.96 |  |
| Hypertension (%) |  |  |  |  |
| Yes | 36.99 | 44.55 | 50.38 | <0.0001 |
| No | 63.01 | 55.45 | 49.62 |  |
| Diabetes (%) |  |  |  | <0.0001 |
| Yes |  |  |  |  |
| No | 7.9 | 11.3 | 19.53 |  |
|  | 92.1 | 88.7 | 80.47 |  |
| BMI (kg/m2) | 26.55 ± 5.30 | 28.82 ± 5.27 | 30.20 ± 5.32 | <0.0001 |
| SBP (mmHg) | 123.44 ± 18.31 | 125.25 ± 16.70 | 127.41 ± 17.73 | <0.0001 |
| DBP (mmHg) | 70.78 ± 10.56 | 71.17 ± 12.72 | 71.62 ± 13.77 | 0.3216 |
| TC (mg/dL) | 97.45 ± 60.27 | 129.28 ± 82.12 | 170.74 ± 115.58 | <0.0001 |
| HbA1c (%) | 5.57 ± 0.85 | 5.70 ± 0.88 | 6.05 ± 1.24 | <0.0001 |
| AST(U/L) | 25.57 ± 19.24 | 25.66 ± 13.60 | 25.12 ± 10.82 | <0.0001 |
| ALT(U/L) | 22.57 ± 15.25 | 25.40 ± 20.59 | 26.25 ± 14.95 | 0.6856 |
| Calcium (mmol/l) | 9.47 ± 0.36 | 9.44 ± 0.34 | 9.45 ± 0.37 | 0.1345 |
| Phosphorus (mmol/L) | 3.86 ± 0.54 | 3.80 ± 0.56 | 3.74 ± 0.58 | <0.0001 |
| Scr (mg/dL) | 0.87 ± 0.38 | 0.92 ± 0.48 | 0.98 ± 0.35 | <0.0001 |
| AAC Scores | 1.16 ± 2.92 | 1.64 ± 3.32 | 2.08 ± 4.10 | <0.0001 |
| AAC(%) | 22.29 | 31.15 | 32.67 | <0.0001 |
| AAC Severe(%) | 5.09 | 7.76 | 10.12 | 0.0001 |

SBP, systolic blood pressure; DBP, diastolic blood pressure; BMI, body mass index; AST, aspartate aminotransferase; ALT, alanine aminotransferase; Scr, serum creatinine; TC, HbA1c, Hemoglobin A1C

Additionally, this group demonstrated a progressive increase in AAC scores (Tertile 1: 1.16 ± 2.92; Tertile2: 1.64 ± 3.32; Tertile3: 2.08 ± 4.10), prevalence of AAC (Tertile 1: 22.29%; Tertile2: 31.15%; Tertile3: 32.67%), and prevalence of severe AAC (Tertile 1: 5.09%; Tertile2: 7.76%; Tertile3: 10.12%).

### Higher MHR Is Associated with a Higher AAC Score

Table 2 show a significant positive correlation was observed between MHR and AAC scores across all models (Model 1: β = 1.53, 95% CI: 1.01,2.06; Model 2: β = 1.36, 95% CI: 0.85,1.87; Model 3: β = 1.22, 95% CI: 0.67, 1.76. In Model 3, after full adjustment, each unit increase in log2-transformed MHR was linked to a 1.22-unit elevation in AAC score. To deepen the analysis of the MHR-AAC score relationship, MHR was categorized into tertiles, transitioning from a continuous to a categorical variable. Within Model 3, comparisons between the Tertile 3 and Tertile 1 revealed a β of 0.90 (95% CI: 0.54, 1.17). The AAC score in Tertile 2 was, on average 0.52 units higher than in Tertile 1, a statistically significant difference.

**Table 2:**
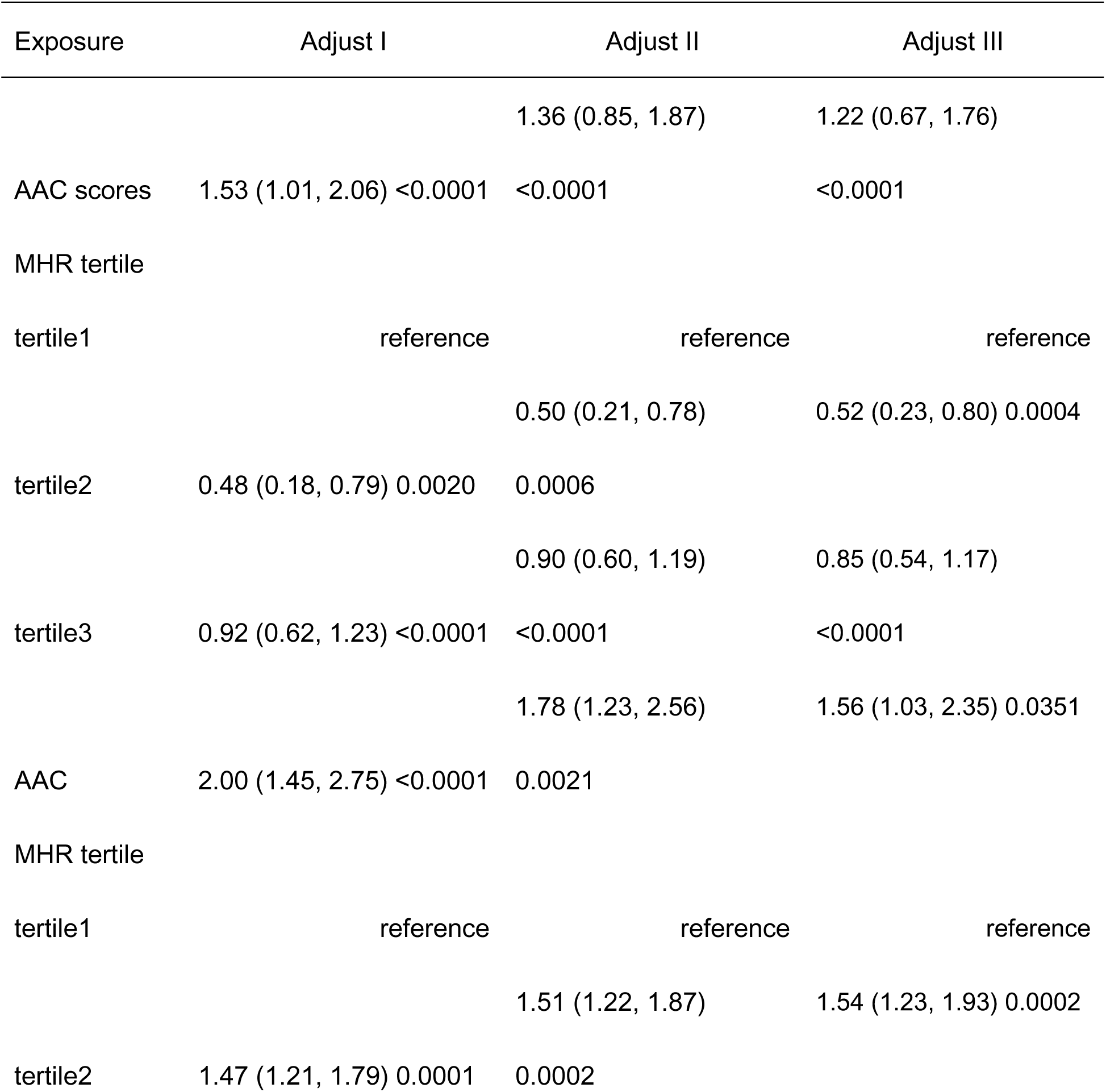
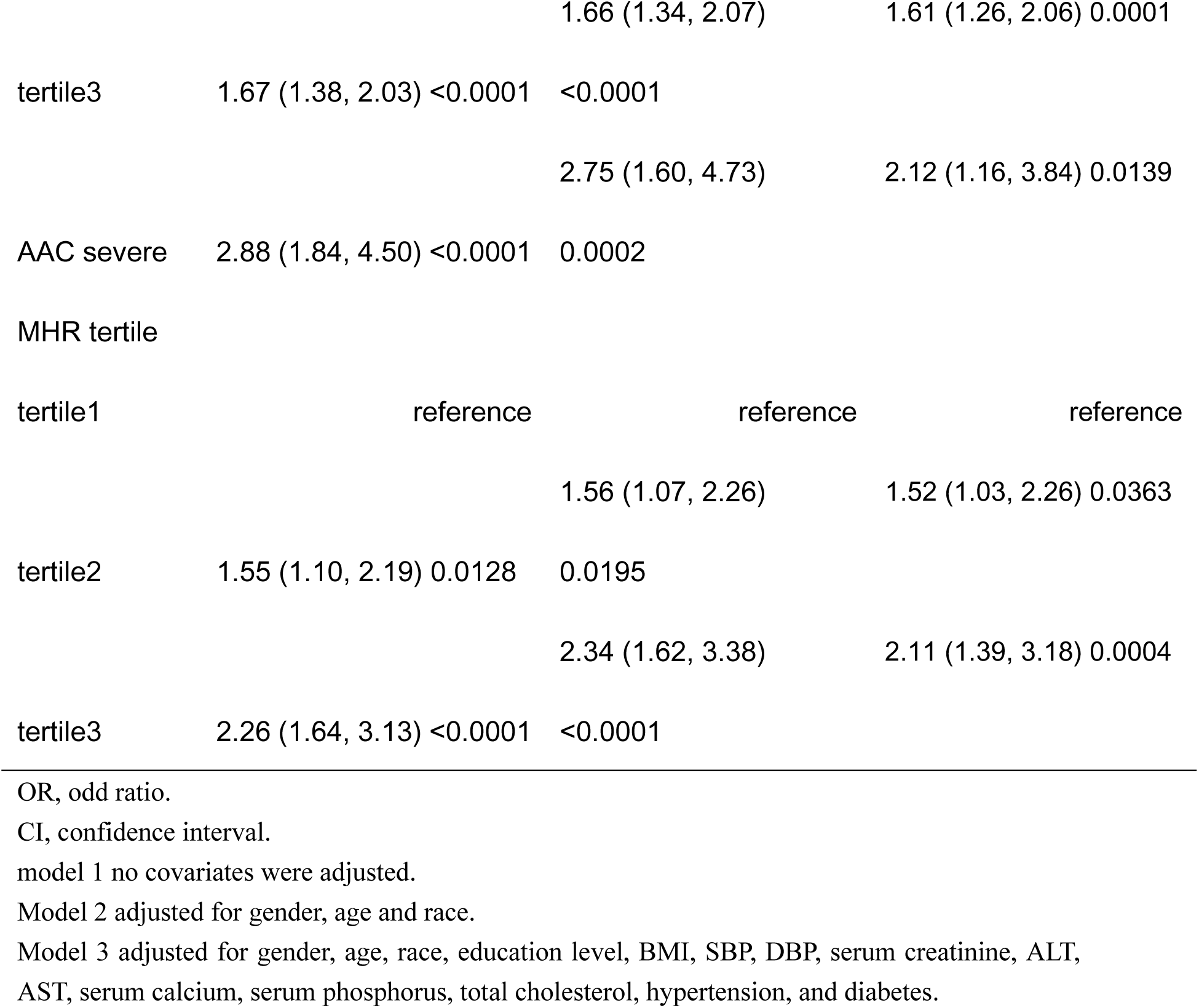
Association of MHR with AAC score, AAC and severe AAC.

### Higher MHR Is Associated with an Increased Risk of AAC and Severe AAC

Our analysis further revealed a significant association between elevated MHR and increased risk for both AAC and severe AAC in table 2. For AAC, odds ratios (ORs) in Model 1, 2, and 3 were 2.00 (95% CI: 1.45, 2.75, p < 0.0001), 1.78 (95% CI: 1.23,2.56, p < 0.002), and 1.56 (95% CI: 1.03, 2.35, p = 0.0351), respectively. For severe AAC, the observed ORs were 2.88 (95% CI: 1.84,4.50, p < 0.0001) in model 1, 2.75 (95% CI: 1.60,4.73, p = 0.0002) in model 2, and 2.12 (95% CI: 1.16, 3.84, p = 0.0139) in model 3. Sensitivity analyses corroborated these findings, with Tertile 3 demonstrating a significant increase in risk for AAC (OR = 1.61, 95% CI: 1.26, 2.06, p = 0.0001) and severe AAC (OR = 2.11, 95% CI: 1.39, 3.18, p=0.0004) relative to Tertile 1, further substantiating the robust positive correlation between elevated MHR and the risk of AAC and severe AAC.

### Associations Between MHR and AAC scores, AAC, severe AAC

Smooth curve fittings based on GAM were conducted to evaluate a possible non-linear dose–response relationship of MHR with AAC scores, AAC, severe AAC. The results showed a non-linear relationship between MHR and AAC score (Figure 2) after adjusting for all covariates. The relationships between MHR and AAC, as well as severe AAC have the same result. The trends were consistent with regressions.

**Figure 2:**
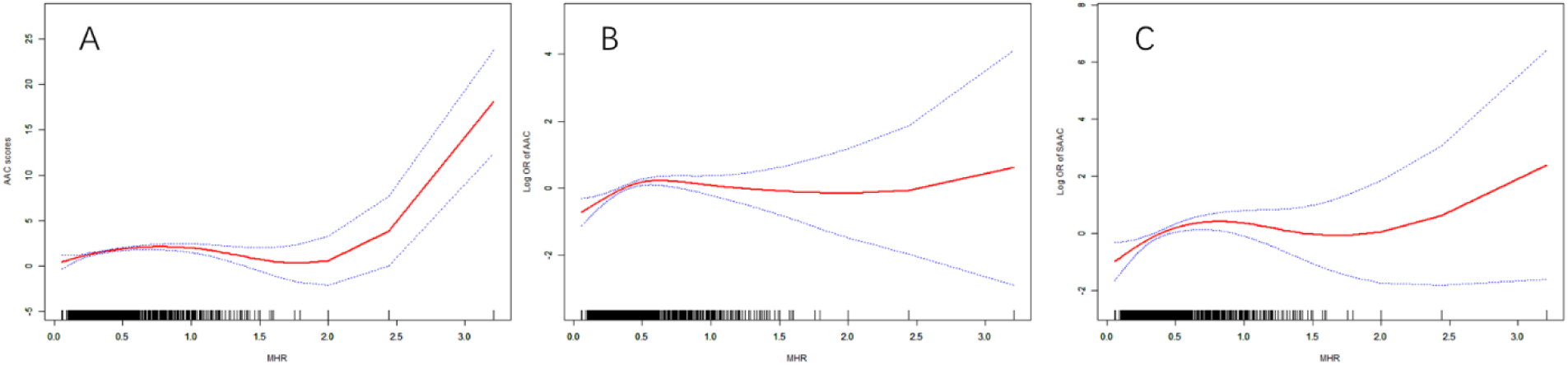
The no-linear dose–response relationship between MHR and AAC score, AAC, SAAC by GAM. (A) The non-linear relationship of HMR with AAC score. (B) The non-linear relationship of HMR with AAC. (C) The non-linear relationship of HMR with SAAC.

### Subgroup Analysis

We conducted subgroup analyses to delve deeper into the relationship between MHR and AAC score, AAC, and severe AAC across various demographics: gender, age, BMI, hypertension, and diabetes in figure (3,4,5). Additionally, interaction tests were carried out to assess the influence of potential effect modifiers on these associations, with p-values for interaction > 0.05 indicating no significant modifier effects. Except for the interaction tests concerning hypertension and BMI with AAC scores, no significant correlation was found (all other p-values for interaction > 0.05), suggesting uniform association magnitudes across varied demographic settings. In subgroups divided by gender, age, BMI, hypertension, and diabetes, significant positive associations were consistently observed between MHR and AAC score, indicating that this relationship holds across different demographic characteristics. Similarly, for both AAC and severe AAC, no significant dependency was found on age, BMI, hypertension, or diabetes status (all p for interaction > 0.05), highlighting a universal positive correlation between MHR and the risk of AAC and severe AAC across diverse populations, making these findings broadly applicable.

**Figure 3.**
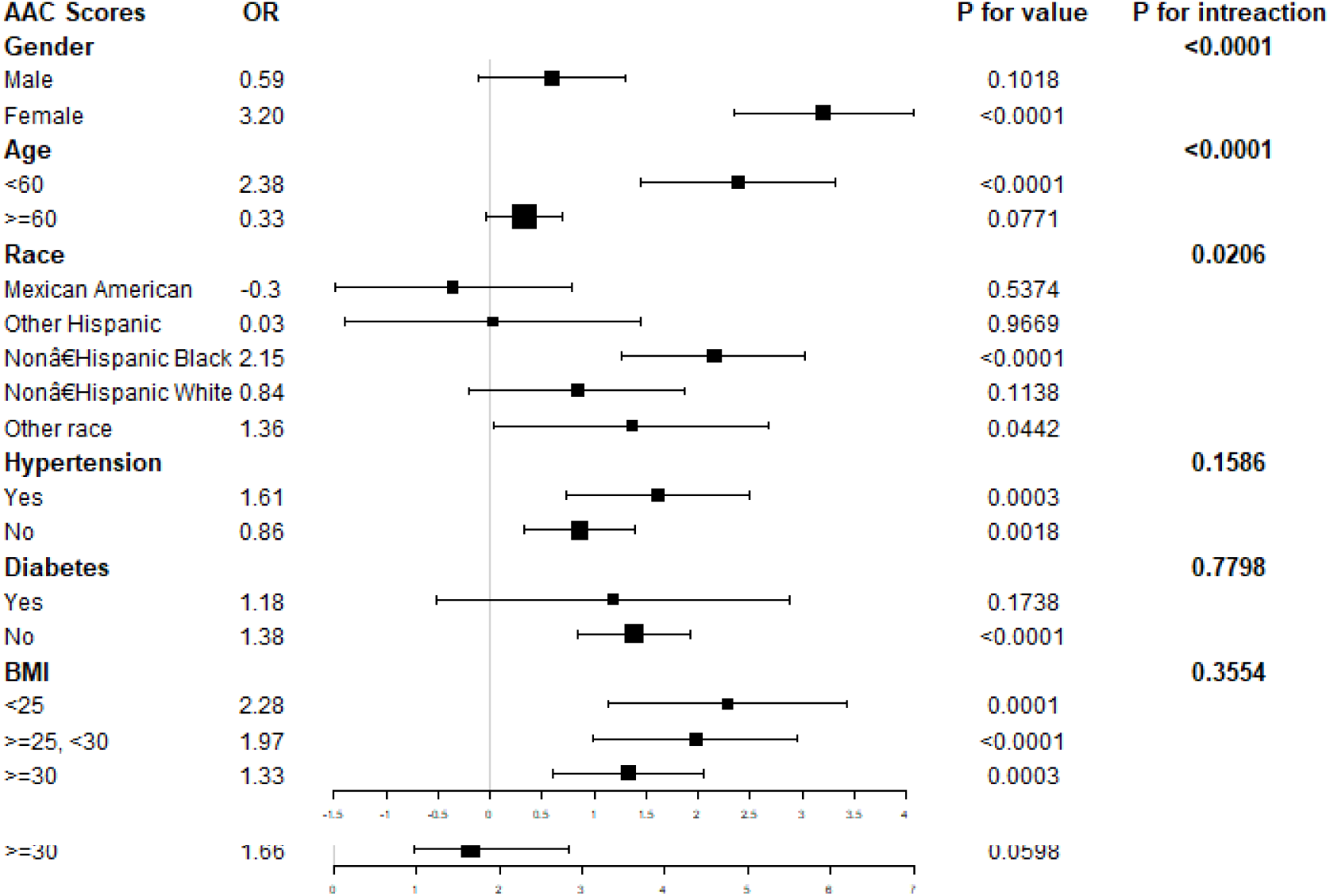
Subgroup analysis for the association between MHR and AAC scores.

**Figure 4.**
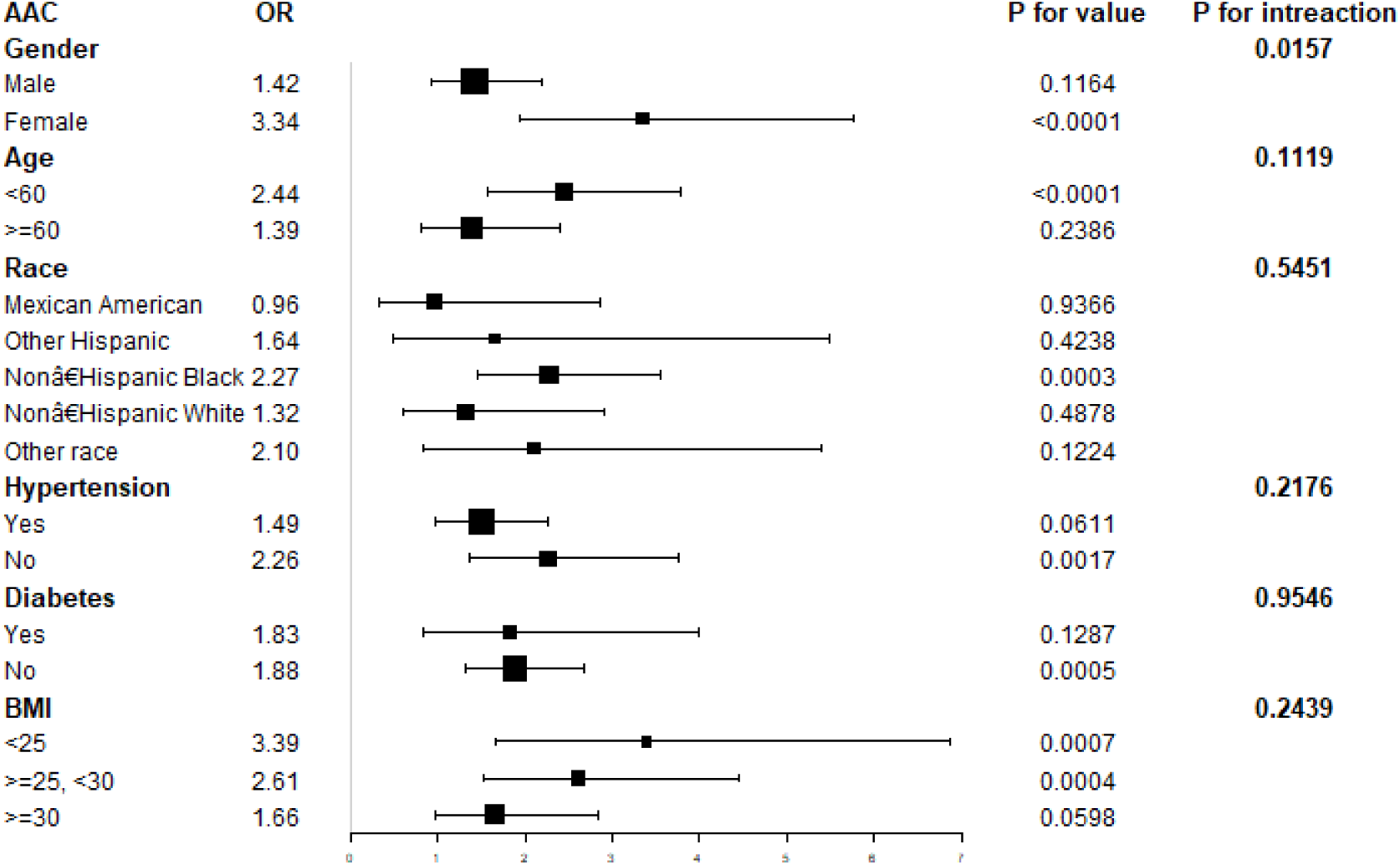
Subgroup analysis for the association between MHR and AAC.

**Figure 5.**
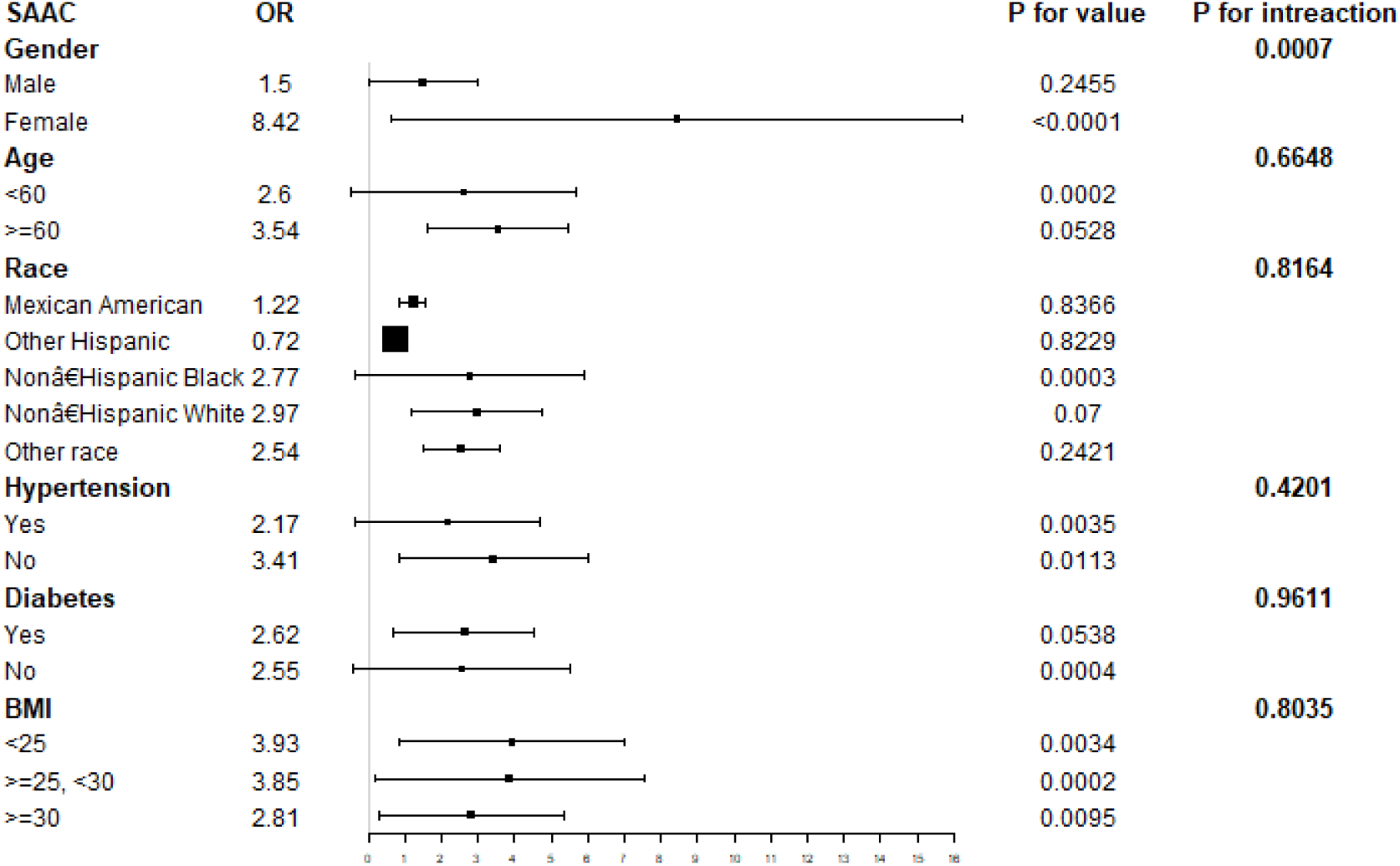
Subgroup analysis for the association between MHR and SAAC.

## Discussion

In this cross-sectional study of 3,017 adults, we found a significant positive correlation between MHR and AAC scores, AAC, and severe AAC. These findings suggest that higher MHR levels may contribute to an increased AAC score and a heightened risk of AAC and severe AAC. Furthermore, the stability of these associations across subgroups stratified by gender, age, BMI, hypertension status, and diabetes status underscores their potential applicability in diverse population settings. This consistency indicates that the observed positive relationship between MHR and AAC risk factors could be relevant across various demographic and health-related spectrums, emphasizing the potential of MHR as a universal marker for AAC risk assessment.

Recent studies have underscored the positive correlation between MHR and cardiovascular diseases, highlighting its potential as a significant marker in assessing the severity of such conditions. Notably, Kundi et al. and Akboga et al. discovered a significant link between MHR and the severity of coronary atherosclerosis, as measured by the SYNTAX score, among patients with stable coronary artery disease[21, 22]. Similarly, BİLGE DURAN KARADUMAN identified MHR as a crucial independent predictor for the progression speed and diagnosis of severe Bicuspid Aortic Valve (BAV) stenosis[23]. Furthermore, Burak Acar demonstrated a significant relationship between MHR and aortic dilatation[24], while Jiani Sun explored the associations between MLR and MHR with AAC in patients undergoing peritoneal dialysis[25]. Our investigation, conducted within the U.S. population, aligns with these findings, reinforcing the notion that MHR can serve as an insightful biomarker for atherosclerosis severity.

Recently, MHR has been increasingly getting more and more attention, suggesting its potential to provide a rapid assessment of atherosclerosis severity through assessment of dyslipidemia and inflammation[11]. Inflammation and dyslipidemia are two important and mutually influential processes in atherosclerosis[26, 27]. Monocytes are the main source of proinflammatory factors during atherosclerosis[28]. The monocyte, being the largest leukocyte in the body, along with its derivatives macrophage and dendritic cells, constitutes an important inflammatory response system in the body. In the early stage, vascular endothelial function is impaired attributable to diverse factors leading to the subsequent release of chemokines such as CC-chemokine ligand 2 into the blood circulation[29]. Through the action of these chemokines, monocytes then adhere firmly to the endothelium, allowing their transmigration into the subendothelial space and differentiation into macrophages and foam cells[30],resulting in smooth muscle proliferation[31]. In contrast to the proatherogenic effects of monocytes, the HDL can slow the progression of atherogenesis. HDL promotes anti-cholesterol transport to reduce the accumulation in arteries[32], hence alleviating the pro-inflammatory and pro-oxidant effects of monocytes[33]. HDL cholesterol prevents the transformation of monocytes into macrophages and the promotion of monocytes’ inflammatory and pro-oxidative effects, combined with inflammatory mediators to neutralize the chemotactic activity of leukocytes and inhibit the aggregation of monocytes. This action inhibits atherosclerosis’ inflammatory processes, thus hindering its progression[34, 35]. HDL protects endothelial cells through a variety of effects, effectively protecting cardiovascular health[36, 37].

One of the strengths of our study is that it utilized nationwide, population-based sampling survey data with appropriate weighting of survey participants, enhancing the findings’ applicability across the US population. Given the association between ACC and various risk factors, several covariates related to AAC, including gender, age, BMI, serum creatinine, hypertension, diabetes, and smoking, were incorporated into the adjusted model for weighted multiple regression analysis. Moreover, the large sample size of our study enabled detailed subgroup analyses. Our findings demonstrate a consistent positive association between MHR and AAC across subpopulations with varying gender, age, BMI, hypertension status, and diabetes status. In the gender-stratified subgroup analysis, MHR exhibited a stronger positive correlation with higher AAC scores (β = 3.20, p for trend < 0.0001) and an increased risk of AAC (OR = 3.34, p for trend < 0.0001) and severe AAC (OR = 8.42, p for trend < 0.0001) among females. The impact of sex hormones may play a pivotal role. Post-menopause, women experience increased cardiovascular risk, a finding supported by previous studies identifying menopause as a female-specific cardiovascular risk factor[38].

The limitations of this study warrant careful consideration. Given that NHANES is a cross-sectional survey of the US population, establishing a causal relationship between dietary inflammatory potential and AAC proves challenging, potentially limiting the generalizability of our findings to broader populations or distinct ethnic groups. Consequently, prospective studies incorporating larger sample sizes are essential to elucidate the causal relationships. Secondly, the presence of residual confounders, attributable to unmeasured or unknown variables, cannot be entirely excluded. For instance, among the non-hospitalized population, medications may influence VC. Furthermore, NHANES 2013–2014 lacked data on AAC scores for participants aged under 40 years, restricting our analysis to broader age groups. The underlying molecular mechanisms supporting our findings remain uncertain, necessitating further laboratory-based experimentation. Accordingly, additional research is imperative to deepen our understanding of the long-term relationship between dietary inflammatory potential and AAC.

## Conclusion

Our study demonstrated that increased MHR is associated with an increased AAC score and a risk of AAC and severe AAC in adults aged ≥ 40 years in the United States, suggesting that MHR exposure has a negative effect on cardiovascular health. However, additional studies are essential to validate our findings.

## Data Availability

All data underpinning the findings presented in this manuscript are derived from the National Health and Nutrition Examination Survey (NHANES) for the years 2013 to 2014. The NHANES is a public domain resource managed by the National Center for Health Statistics (NCHS), which conducts these surveys in 2-year cycles to assess the health and nutritional status of the non-institutionalized U.S. population. As such, the data utilized for our study are publicly accessible and can be obtained directly from the NHANES website at https://www.cdc.gov/nchs/nhanes/. This includes comprehensive datasets on abdominal artery calcification, dietary inflammatory index, and other relevant health metrics considered in our analysis. Our study's specific data subset, criteria for inclusion, and the analytical approach are detailed within the Methods section of our manuscript for reproducibility and transparency.

## Reference

1. Ngai, D., M. Lino, and M.P. Bendeck, Cell-Matrix Interactions and Matricrine Signaling in the Pathogenesis of Vascular Calcification. Frontiers in cardiovascular medicine, 2018. Vol.5: p. 174.

2. Yahagi, K., et al., Pathology of Human Coronary and Carotid Artery Atherosclerosis and Vascular Calcification in Diabetes Mellitus. Arteriosclerosis, Thrombosis & Vascular Biology, 2017. Vol.37(No.2): p. 191–204.

3. NJ, N.J.P.P. and C.M.G.G. CM, A current understanding of vascular calcification in CKD. Am J Physiol Renal Physiol, 2014. Vol.307(No.8): p. F891-F900.

4. Zhu, J., et al., Relationship between carotid or coronary artery calcification and osteoporosis in the elderly. Minerva medica, 2019. Vol.110(No.1): p. 12–17.

5. Mäkelä, S., et al., Abdominal Aortic Calcifications Predict Survival in Peritoneal Dialysis Patients. Peritoneal Dialysis International, 2018. Vol.38(No.5): p. 366–373.

6. Bartstra, J.W., et al., Abdominal aortic calcification: from ancient friend to modern foe. European journal of preventive cardiology, 2021. Vol.28(No.12): p. 1386–1391.

7. Hendriks, E.J.E., et al., Annularity of Aorto-Iliac Arterial Calcification and Risk of All-Cause and Cardiovascular Mortality. JACC. Cardiovascular imaging, 2018. Vol.11(No.11): p. 1718–1719.

8. SH, S., et al., Abdominal Aortic Calcification and Cardiovascular Outcomes in Chronic Kidney Disease: Findings from KNOW-CKD Study. Journal of clinical medicine, 2022. Vol.11(No.5): p. 1157.

9. Kevin, L., et al., Prognostic Value of Abdominal Aortic Calcification: A Systematic Review and Meta-Analysis of Observational Studies. Journal of the American Heart Association [electronic resource], 2021. Vol.10(No.2): p. e017205.

10. Villanueva, D.L.E., et al., Monocyte to High-Density Lipoprotein Ratio (MHR) as a predictor of mortality and Major Adverse Cardiovascular Events (MACE) among ST Elevation Myocardial Infarction (STEMI) patients undergoing primary percutaneous coronary intervention: a meta-analysis. Lipids in Health & Disease, 2020. Vol.19(No.1): p. 1–8.

11. Kanbay, M., et al., Monocyte count/HDL cholesterol ratio and cardiovascular events in patients with chronic kidney disease. International Urology and Nephrology, 2014. Vol.46(No.8): p. 1619–1625.

12. Chen, J., et al., The Role of Monocyte to High-Density Lipoprotein Cholesterol Ratio in Prediction of Carotid Intima-Media Thickness in Patients With Type 2 Diabetes. Frontiers in endocrinology, 2019. Vol.10: p. 191.

13. Dyah Samti Mayasari, N.T., Hariadi Hariawan, Association of monocyte-to-high density lipoprotein ratio with arterial stiffness in patients with diabetes. BMC cardiovascular disorders, 2021. Vol.21(No.1): p. 362.

14. Wang, H.-Y., et al., Assessing the performance of monocyte to high-density lipoprotein ratio for predicting ischemic stroke: insights from a population-based Chinese cohort. Lipids in Health and Disease, 2019. Vol.18(No.1): p. 1–11.

15. Zhan, X., et al., Monocyte to high-density lipoprotein ratio and cardiovascular events in patients on peritoneal dialysis. Nutrition, metabolism, and cardiovascular diseases : NMCD, 2020. Vol.30(No.7): p. 1130–1136.

16. Skubisz C, K.A., National Health and Nutrition Examination Survey. Plan and, Aps Meeting., 2013.

17. Xiao-Cong Liu, G.-D.H., Kenneth Lo, Yu-Qing Huang, Ying-Qing Feng, The Triglyceride-Glucose Index, an Insulin Resistance Marker, Was Non-linear Associated With All-Cause and Cardiovascular Mortality in the General Population. Frontiers in cardiovascular medicine, 2021. Vol.7: p. 628109.

18. Kauppila, L., New indices to classify location, severity and progression of calcific lesions in the abdominal aorta: a 25-year follow-up study. Atherosclerosis, 1997. Vol.132(No.2): p. 245–250.

19. Bandeira, E., et al., Association Between Vascular Calcification and Osteoporosis in Men With Type 2 Diabetes(Article). Journal of Clinical Densitometry, 2012. Vol.15(No.1): p. 55–60.

20. W, C., et al., Association between dietary zinc intake and abdominal aortic calcification in US adults. Nephrology, dialysis, transplantation : official publication of the European Dialysis and Transplant Association - European Renal Association, 2020. Vol.35(No.7): p. 1171–1178.

21. Akboga, M.K., et al., Usefulness of monocyte to HDL-cholesterol ratio to predict high SYNTAX score in patients with stable coronary artery disease. Biomarkers in medicine, 2016. Vol.10(No.4): p. 375–383.

22. H, K., et al., Association of monocyte/HDL-C ratio with SYNTAX scores in patients with stable coronary artery disease. HERZ, 2016. Vol.41(No.6): p. 523–529.

23. Bİlge Duran Karaduman, H.A., Telat KeleŞ, Engİn Bozkurt, Association between monocyte-to-high density lipoprotein-cholesterol ratio and bicuspid aortic valve degeneration. Turkish journal of medical sciences, 2020. Vol.50(No.5): p. 1307–1313.

24. Acar, B., et al., Monocyte-to-HDL-cholesterol ratio is associated with Ascending Aorta Dilatation in Patients with Bicuspid Aortic Valve. African Health Sciences, 2021. Vol.21(No.1): p. 96–104.

25. Jiani Sun, Y.L., Lei Shen, Deyu Xu, Wengang Sha, Ling Zhou, Jianzhong Li, Clinical study on the correlation between monocyte-related ratios and calcification of the abdominal aorta in peritoneal dialysis patients. Therapeutic apheresis and dialysis : official peer-reviewed journal of the International Society for Apheresis, the Japanese Society for Apheresis, the Japanese Society for Dialysis Therapy, 2023. Vol.27(No.4): p. 742–751.

26. Libby, P., et al., Progress and challenges in translating the biology of atherosclerosis. Nature, 2011. Vol.473(No.7347): p. 317–325.

27. Lucchi, T.A., Dyslipidemia and prevention of atherosclerotic cardiovascular disease in the elderly. Minerva Medica, 2022. Vol.112(No.6): p. 804–816.

28. Swirski, F.K. and M. Nahrendorf, Leukocyte Behavior in Atherosclerosis, Myocardial Infarction, and Heart Failure. Science, 2013. Vol.339(No.6116): p. 161–166.

29. Yang, J., et al., Role of MCP-1 in tumor necrosis factor-alpha-induced endothelial dysfunction in type 2 diabetic mice. American journal of physiology. Heart and circulatory physiology, 2009. Vol.297(No.4): p. H1208–H1216.

30. Ann-Kathrin Vlacil, J.S., Bernhard Schieffer, Karsten Grote, Variety Matters: Diverse Functions of Monocyte Subtypes in Vascular Inflammation and Atherogenesis. Vascular pharmacology, 2019. Vol.113(No.1): p. 9–19.

31. Moreno, P.R., et al., Intomomedial interface damage and adventitial inflammation is increased beneath disrupted atherosclerosis in the aorta: Implications for plaque vulnerability. Circulation, 2002. Vol.105(No.21): p. 2504–2511.

32. Rosenson, R.S., et al., Dysfunctional HDL and atherosclerotic cardiovascular disease. Nature Reviews Cardiology, 2015. Vol.13(No.1): p. 48–60.

33. Murphy, A.J., et al., *High-density lipoprotein reduces the human monocyte inflammatory response.* Arteriosclerosis, Thrombosis, and Vascular Biology, 2008. Vol.28(No.11): p. 2071–2077.

34. Yvan-Charvet, L., et al., ATP-Binding Cassette Transporters and HDL Suppress Hematopoietic Stem Cell Proliferation. Science, 2010. Vol.328(No.5986): p. 1689–1693.

35. Di Bartolo, B.A., et al., Infusional high-density lipoproteins therapies as a novel strategy for treating atherosclerosis. Archives of Medical Science, 2017. Vol.13(No.1): p. 210–214.

36. Ucar, F.M., A potential marker of bare metal stent restenosis: monocyte count - to-HDL cholesterol ratio. BMC Cardiovascular Disorders, 2016. Vol.16(No.1): p. 186.

37. Honigberg, M.C.Z., Seyedeh Maryam Aragam, Krishna Finneran, Phoebe Klarin, Derek Bhatt, Deepak L. Januzzi, James L. Scott, Nandita S. Natarajan, Pradeep Januzzi, James L Jr, Association of Premature Natural and Surgical Menopause With Incident Cardiovascular Disease. JAMA: Journal of the American Medical Association, 2019. Vol.322(No.24): p. 2411–2421.

38. Appelman, Y., et al., Sex differences in cardiovascular risk factors and disease prevention. Atherosclerosis, 2015. Vol.241(No.1): p. 211–218.

